# Impaired cellular immunity to SARS-CoV-2 in severe COVID-19 patients

**DOI:** 10.1101/2020.08.10.20171371

**Authors:** Ling Ni, Meng-Li Cheng, Hui Zhao, Yu Feng, Jingyuan Liu, Fang Ye, Qing Ye, Gengzhen Zhu, Xiaoli Li, Pengzhi Wang, Jing Shao, Yong-Qiang Deng, Peng Wei, Fang Chen, Cheng-Feng Qin, Guoqing Wang, Fan Li, Hui Zeng, Chen Dong

**Affiliations:** Institute for Immunology and School of Medicine, Tsinghua University, Beijing 100084, China; Center for Human Disease Immuno-monitoring, Beijing Friendship Hospital, Beijing 100050, China; College of Basic Medical Science, Jilin University, Changchun 130021, China; Department of Virology, State Key Laboratory of Pathogen and Biosecurity, Institute of Microbiology and Epidemiology, Academy of Military Medical Sciences, Beijing 100071, China; Beijing Ditan Hospital, Capital Medical University, and Beijing Key Laboratory of Emerging Infectious Diseases, Beijing100015, China; Department of Hematology, Chui Yang Liu Hospital affiliated to Tsinghua University, Beijing 100022, China; Department of Cardiology, Chui Yang Liu Hospital affiliated to Tsinghua University, Beijing100022, China; China-Japan Union Hospital, Jilin University, Changchun 130033, China; Beijing Key Lab for Immunological Research on Chronic Diseases, Beijing 100084, China

**Keywords:** SARS-CoV-2, acute respiratory distress syndrome, adaptive immunity, neutralization antibody, T cells

## Abstract

The World Health Organization has declared SARS-CoV-2 virus outbreak a world-wide pandemic. Individuals infected by the virus exhibited different degrees of symptoms, the basis of which remains largely unclear. Currently, though convalescent individuals have been shown with both cellular and humoral immune responses, there is very limited understanding on the immune responses, especially adaptive immune responses, in patients with severe COVID-19. Here, we examined 10 blood samples from COVID-19 patients with acute respiratory distress syndrome (ARDS). The majority of them (70%) mounted SARS-CoV-2-specific humoral immunity with production of neutralizing antibodies. However, compared to healthy controls, the percentages and absolute numbers of both NK cells and CD8^+^ T cells were significantly reduced, accompanied with decreased IFNγ expression in CD4^+^ T cells in peripheral blood from severe patients. Most notably, we failed in detecting SARS-CoV-2-specific IFNγ production by peripheral blood lymphocytes from these patients. Our work thus indicates that COVID-19 patients with severe symptoms are associated with defective cellular immunity, which not only provides insights on understanding the pathogenesis of COVID-19, but also has implications in developing an effective vaccine to SARS-CoV-2.

## Introduction

Identified in December, 2019, coronavirus disease 2019 (COVID-19) caused by severe acute respiratory syndrome coronavirus 2 (SARS-CoV-2) (Wang et al., 2020a) has become a global public health threat. As of July 17th, 2020, 13,378,853 global cases were confirmed, of which 580,045 patients died (https://www.who.int/emergencies/diseases/novel-coronavirus-2019/situation-reports). The high infection rate and rapid spread make it a world-wide emergency (Di Pierro et al., 2020). However, up to date, there is no specific anti-viral medicine or vaccine available to prevent or treat COVID-19 and the current standard care relies on supportive treatments.

Approximately 80% of COVID-19 cases are asymptomatic or manifest mild symptoms resembling simple upper respiration tract infection. The remaining COVID-19 patients show severe or critical symptoms with severe pneumonia and acute respiratory distress syndrome (ARDS) (Gruszecka and Filip, 2020). How the immune system modifies and controls the infection is still not well understood, which may underscore the different degrees of symptoms amongst the population. Several papers were reported on the adaptive immune responses in the recovered mild COVID-19 patients. We detected SARS-CoV-2-specific antibodies and T cells in convalescent individuals (Ni et al., 2020). Grofoni et al also found that ~70% and 100% of COVID-19 convalescent subjects with mild and severe symptoms developed SARS-CoV-2-specific CD8^+^ and CD4^+^ T cells, respectively (Grifoni et al., 2020). However, there is very limited understanding on immune responses in severe COVID-19 patients during hospitalization. In a retrospective study, Chen et al observed that absolute numbers of T lymphocytes, both CD4^+^ and CD8^+^, were markedly lower in severe patients than those in moderate cases (Chen et al., 2020). In contrast to the aforementioned study, Zheng et al did not observe reduced lymphocytes in the severe disease group, but T cells showed elevated exhaustion levels and reduced functional heterogeneity (Zheng et al., 2020). Using megapools of overlapping or prediction-based peptides covering the SARS-CoV-2 proteome, Weiskopf et al found CD4^+^ and CD8^+^ T cells showed activation marker expression in 10 out of 10 and 8 out of 10 patients with severe COVID-19 during hospitalization, respectively (Weiskopf et al., 2020). For humoral responses, Lynch et al found that antibody responses were higher in patients with severe than mild disease (Lynch et al., 2020). And high levels of neutralizing antibodies was induced 10 days post onset in both mild and severe patient, which were higher in severe group (Wang et al., 2020b).

In order to understand immune responses and the mechanism underlying the pathogenesis of severe COVID-19, we collected blood samples from 10 patients with ARDS, analyzed their cellular and humoral responses specific to SARS-CoV-2, and compared severe and convalescent patients. Our data reveal defective cellular immunity associated with severe COVID-19. This study provides new insights on the precise treatment of COVID-19 patients and evaluation of candidate vaccines.

## Results

### Detection of SARS-CoV-2-specific antibodies in severe COVID-19 subjects

To understand the immune responses to SARS-CoV-2 in severe patients, we studied 10 patients with acute respiratory distress syndrome (ARDS). Their clinical and pathological characteristics are shown in Table 1. All the patients were hospitalized at Beijing Ditan Hospital and showed severe symptoms via CT scan and were positive in SARS-CoV-2 nucleic acid testing. The mean age was 57.5 years and half of them were female. Among them, 8 (80%) showed lymphopenia. As of today, 1 patient (Pt#9) passed away and the remaining ones had recovered and were discharged from hospital. The blood samples were obtained within 20 days post symptom onset and the detail sampling day for each patient was also shown in Table 1. Human AB serum collected from healthy male AB donors in the US (GemCell, CA) was used as a negative control. Additionally, sera from nine healthy donors were obtained before the SARS-CoV-2 outbreak (HD#1-9). 5 additional healthy donors (HD#10-14) without SARS-CoV-2 infection were analyzed in our T cell assays.

**Table 1.**
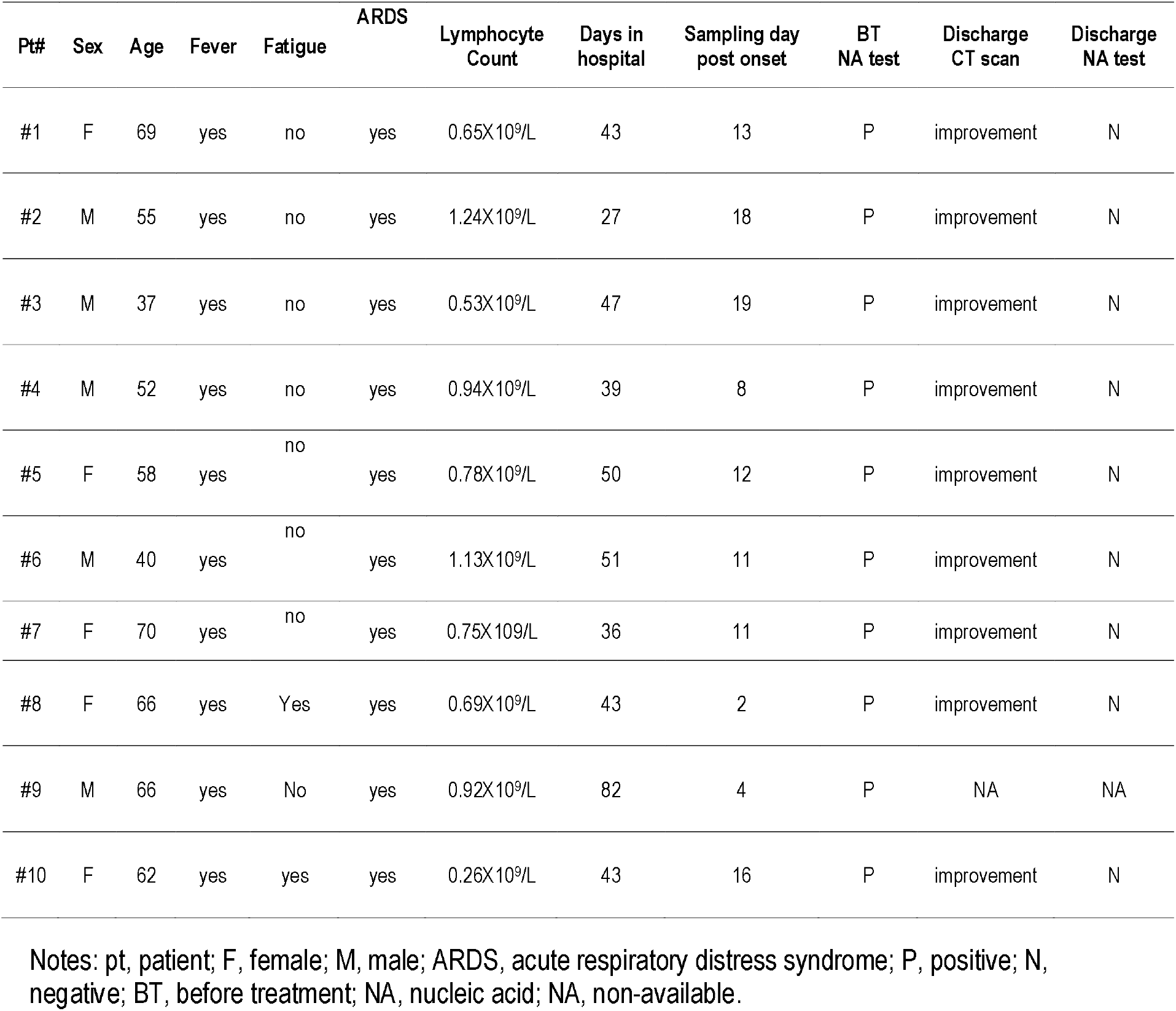
Clinical and pathological characteristics of the severe COVID-19 patients.

| Pt# | Sex | Age | Fever | Fatigue | ARDS | Lymphocyte Count | Days in hospital | Sampling day post onset | BT NA test | Discharge CT scan | Discharge NA test |
| --- | --- | --- | --- | --- | --- | --- | --- | --- | --- | --- | --- |
| #1 | F | 69 | yes | no | yes | 0.65X10<sup>9</sup>/L | 43 | 13 | P | improvement | N |
| #2 | M | 55 | yes | no | yes | 1.24X10<sup>9</sup>/L | 27 | 18 | P | improvement | N |
| #3 | M | 37 | yes | no | yes | 0.53X10<sup>9</sup>/L | 47 | 19 | P | improvement | N |
| #4 | M | 52 | yes | no | yes | 0.94X10<sup>9</sup>/L | 39 | 8 | P | improvement | N |
| #5 | F | 58 | yes | no | yes | 0.78X10<sup>9</sup>/L | 50 | 12 | P | improvement | N |
| #6 | M | 40 | yes | no | yes | 1.13X10<sup>9</sup>/L | 51 | 11 | P | improvement | N |
| #7 | F | 70 | yes | no | yes | 0.75X10<sup>9</sup>/L | 36 | 11 | P | improvement | N |
| #8 | F | 66 | yes | Yes | yes | 0.69X10<sup>9</sup>/L | 43 | 2 | P | improvement | N |
| #9 | M | 66 | yes | No | yes | 0.92X10<sup>9</sup>/L | 82 | 4 | P | NA | NA |
| #10 | F | 62 | yes | yes | yes | 0.26X10<sup>9</sup>/L | 43 | 16 | P | improvement | N |

Notes: pt, patient; F, female; M, male; ARDS, acute respiratory distress syndrome; P, positive; N, negative; BT, before treatment; NA, nucleic acid; NA, non-available.

Using sera from patients and healthy donors, IgG and IgM specific to SARS-CoV-2 NP and S-RBD antigens were analyzed using ELISA assay previously reported (Ni et al., 2020). The individual serum samples were performed by serial dilutions to calculate the area under the curve (AUC) values (Figure 1A). Compared with healthy donors, patients with severe COVID-19 showed significantly elevated anti-NP IgG AUC values (Figure 1B). The AUC values of anti-S-RBD IgG in severe cases were also significantly increased compared to those in healthy controls. Serum NP- and S-RBD-specific IgM antibodies showed significantly higher AUC values in severe COVID-19 patients than in healthy controls (Figure 1B). Notably, patients #1, 4 and 7 did not develop NP- and S-RBD -specific antibody responses, including IgM and IgG. As shown in Figure 1C, anti-NP and S-RBD IgG in severe patients was mainly IgG1 isotype, as in convalescent individuals (Ni et al., 2020). We did not detect IgG2 to either NP or S-RBD proteins (data not shown).

**Figure 1.**
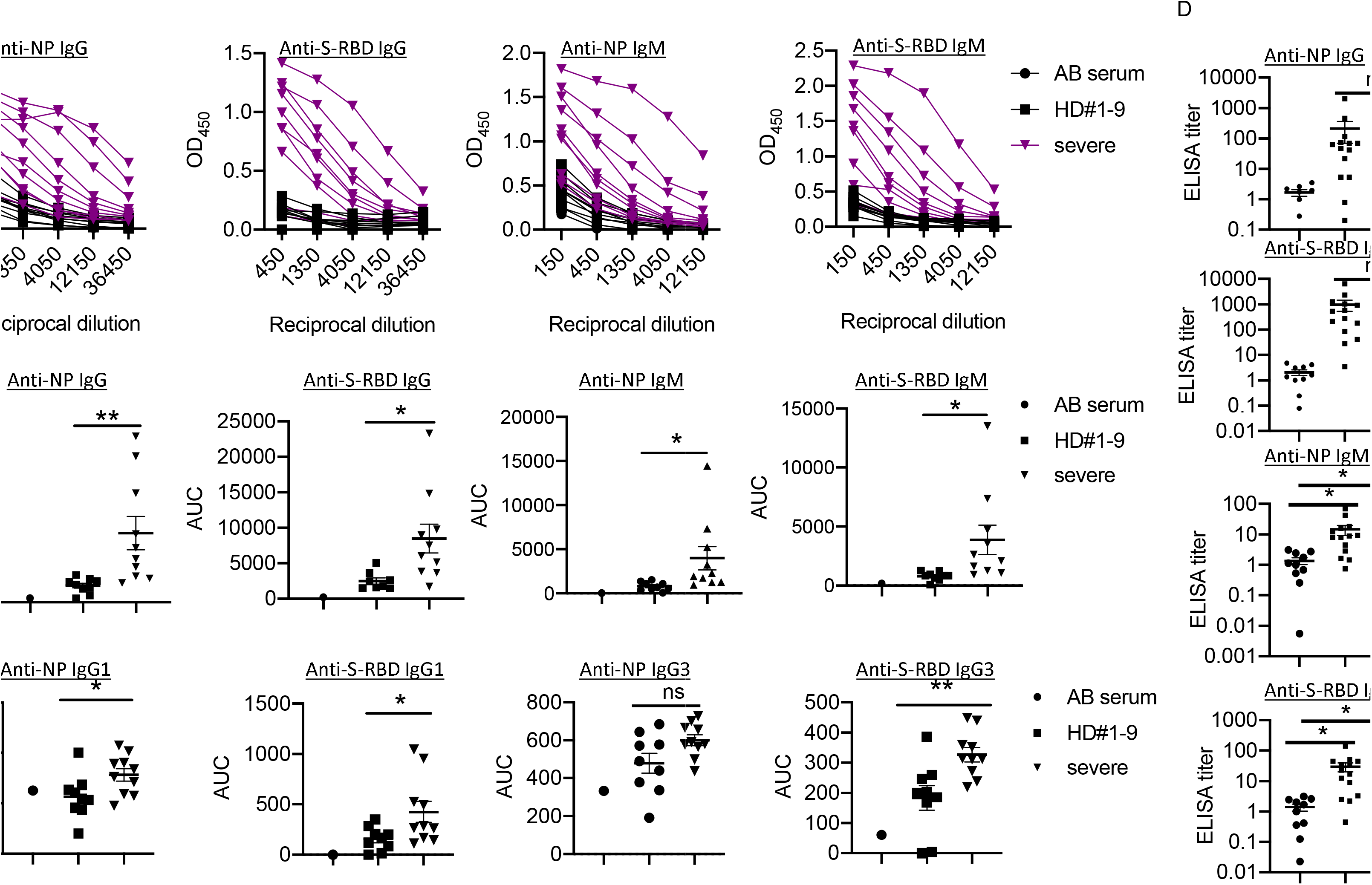
Presence of SARS-CoV-2 NP- and S-RBD-specific antibodies in severe COVID-19 patients. (A) Titration of individual serum samples. (B) Data from the same experiments as (A) were presented as area under curve (AUC). (C) IgG isotypes of 10 COVID-19 patients to recombinant NP and S-RBD. (D) Serum ELISA titers to NP and S-RBD in sera from convalescent individuals (n=14) and severe patients (n=10). The experiment was performed in duplicates. Date are presented as Mean ± SEM. NP, nucleocapsid protein. S-RBD, receptor binding domain of spike protein. HD, healthy donor. Pt, patient. AUC, area under curve. HD#1-9, the sera were collected before SARS-CoV-2 outbreak. *P<0.05, 0.05<**P<0.001, ***P<0.001. ns, not significant.

In order to understand the pathogenic mechanisms in severe patients, we compared the levels of virus-specific IgG or IgM in these patients with those in convalescent individuals. Serum from one convalescent COVID-19 patient was used as a positive control standard to calculate the antibody titers (relative units) for all samples using non-linear regression interpolations (Grifoni et al., 2020). Of note, we observed no significant differences in anti-NP/S-RBD IgG or IgM between these two groups (Figure 1D).

Taken together, these findings indicate that most severe COVID-19 patients mounted IgG and IgM responses specific to SARS-CoV-2 proteins, NP and S-RBD. However, the levels of humoral immune responses varied among the patients.

### Measurement of neutralizing antibody production in severe COVID-19 subjects

To determine the neutralization capacity in sera from patients with severe COVID-19, we performed pseudovirus particle-based neutralization assay as previously described (Ni et al., 2020). As shown in Figure 2A and 2B, patients #1, 4 and 7, did not produce neutralizing antibodies, while patient #3 had a high neutralizing antibody titer (NAT50>1000). Most severe patients (70%) had protective humoral immunity to SARS-CoV-2. Notably, sera from severe patients did not show significantly reduced NAT50 values than convalescent sera (Figure 2C). About 30% of severe patients and 8% of convalescent individuals did not produce neutralizing antibodies, respectively (Figure 2D). Around 40%, 20% and 10% of severe patients showed low (NAT50: 30-500), medium (NAT50: 500-1000) and high (NAT50: >1000) NAT50, respectively, while 50%, 21% and 21% in the convalescent group did.

**Figure 2.**
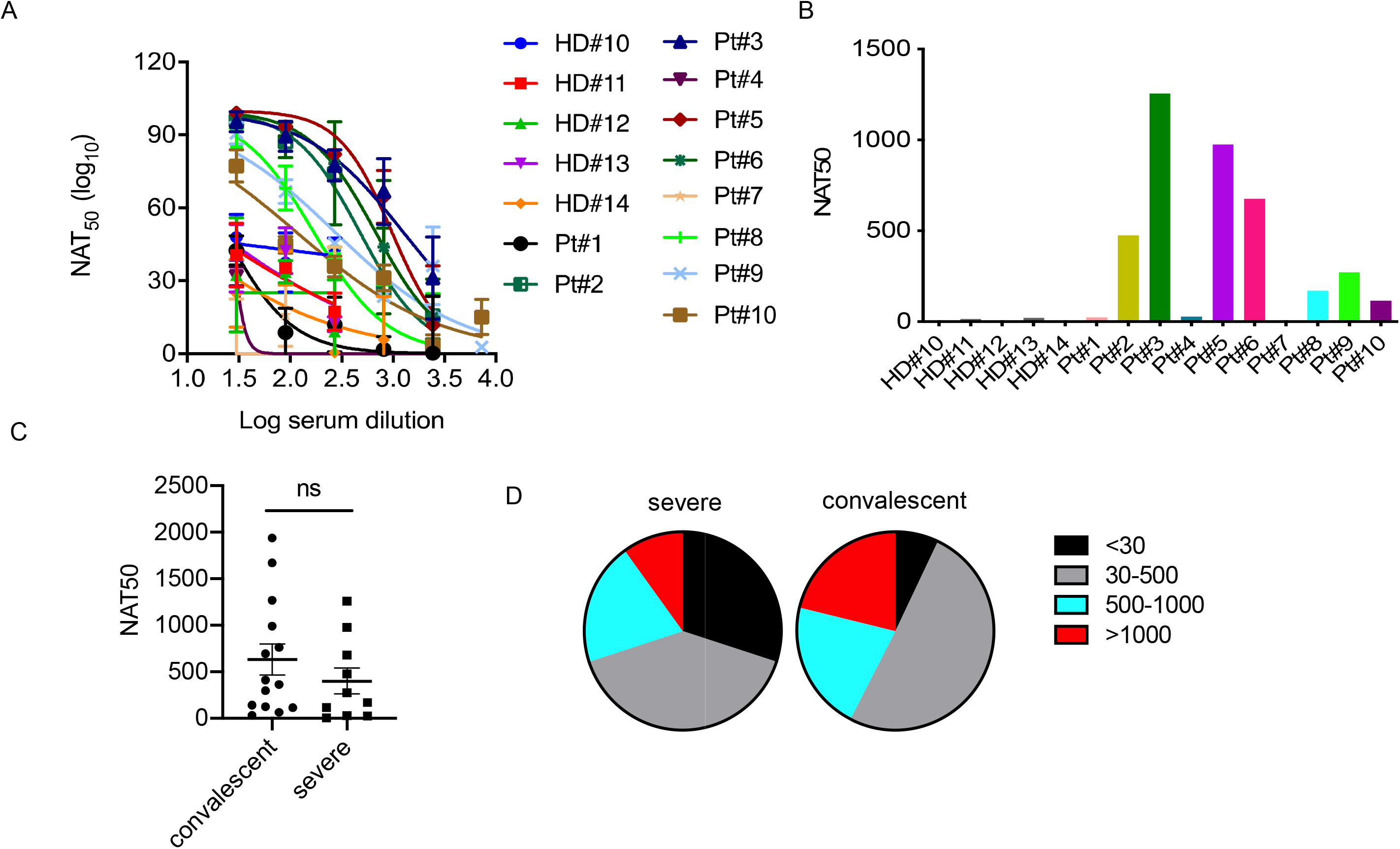
Measurement of neutralizing antibody titers in severe COVID-19 cases. (A) Neutralizing curves of 10 COVID-19 patients measured by pseudovirus-based assay. The experiment with patients was performed in duplicates. (B) Neutralizing antibody titers of 10 severe COVID-19 patients. (C) Comparison of NAT50 between convalescent subjects (n=14) and severe patients (n=10). Date are presented as Mean ± SEM. (D) Pie plot showing NAT50 range in severe and convalescent patients. HD, healthy donor (HD#10-14). Pt, patient. NAT50, neutralizing antibody titers. *P<0.05, 0.05<**P<0.001, ***P<0.001. ns, not significant.

Nonetheless, our results indicate that most severe patients had serum neutralizing antibodies to SARS-CoV-2 infection.

### Lymphocyte numbers and function in severe COVID-19 subjects

To analyze cellular immune responses to SARS-CoV-2, peripheral blood mononuclear cells (PBMCs) from 10 patients with severe COVID-19 and 5 healthy donors were phenotypically analyzed by flow cytometry (Figure 3A). We did not detect live PBMCs in 3 out of 10 patients (Pts# 3, 4 and 5), possibly due to technical issues during cryopreservation. Compared to healthy donors, there was a significant decrease in the percentages of NK cells in the severe patients, but similar frequencies of NKT cells (Figure 3B). Although there was a trend towards increased frequencies of T cells (CD3^+^CD56^-^) in the patient blood, the absolute numbers of T cells were significantly decreased compared to those in blood from healthy controls (Figure 3C). Notably, the percentages and absolute numbers of CD8^+^ T cells were reduced dramatically, while the frequency of CD4^+^ T cells was elevated compared with those in healthy donors. As a result, the CD4:CD8 ratios were significantly higher in severe patients than in the healthy donors (6.7 ± 1.3 in COVID-19 vs 2.5 ± 0.3 in HD, P=0.0226) (Figure 3B). Thus, the severe patients exhibited reduced cytotoxic lymphocytes, both NK and CD8^+^ T cells.

**Figure 3.**
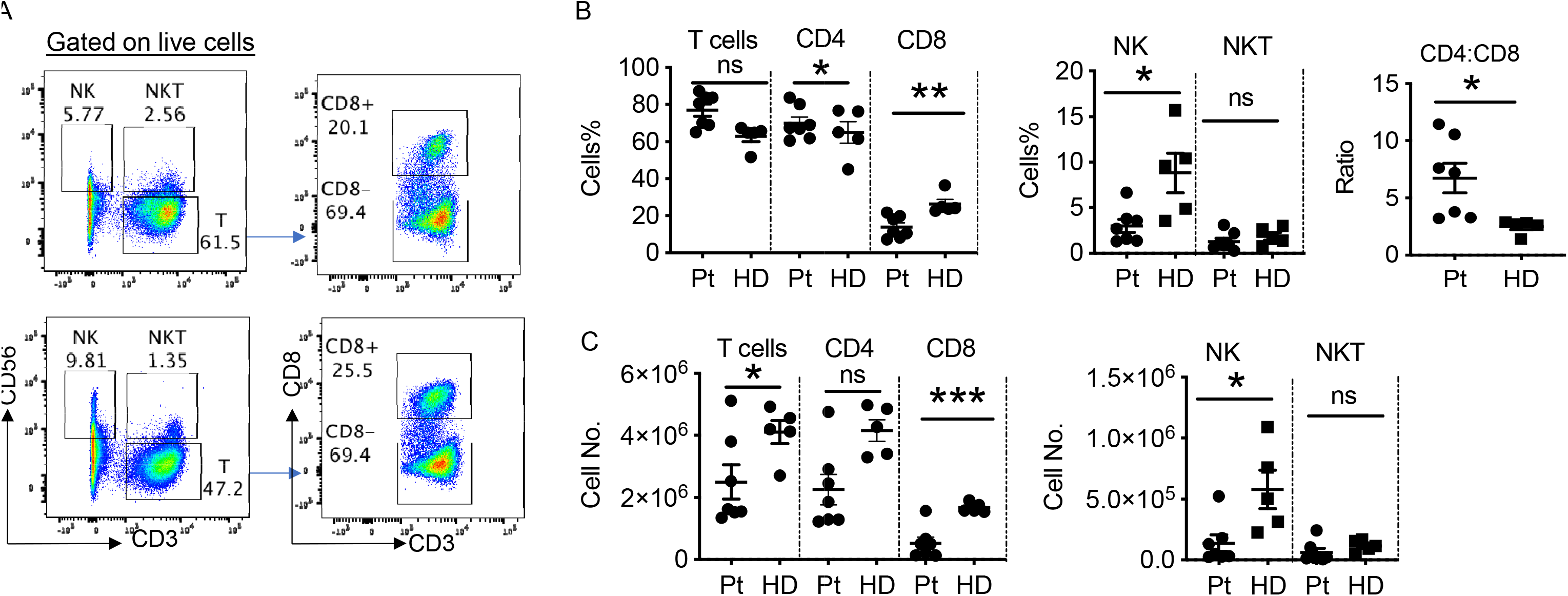
Phenotypic analysis of blood lymphocytes in severe COVID-19 patients. (A) Phenotypic analysis of PBMCs from one representative COVID-19 patient. (B) Summarized data on the frequencies of different immune cell subsets in COVID-19 patients and the ratio of CD4:CD8 T cells. (C) Summarized data on the absolute numbers of different immune cell subsets in COVID-19 patients. HD, healthy donors (HD#10-14); Pt, patients (n=7). Date are presented as Mean ± SEM. *P<0.05, 0.05<**P<0.001, ***P<0.001. ns, not significant.

To further assess the function of the T cells in the severe patients, we stimulated T cells with phorbol myristate acetate (PMA) and ionomycin and then measured cytokine production. As shown in Figure 4A and 4B, CD4^+^ T cells from severe patients expressed significantly lower levels of IFNγ than those from healthy donors. We observed no apparent difference in the frequencies of TNFα-expressing CD4^+^ T cells, but significantly reduced percentages of IFNγ^+^ TNFα^+^ T cells (Figure 4C and 4D), consistent with a published report (Chen et al., 2020). Only 2 out of 7 patients had detectable IL-17A^+^ CD4^+^ T cells, which expressed TNFα, but not IFNγ (Figure 4D).

**Figure 4.**
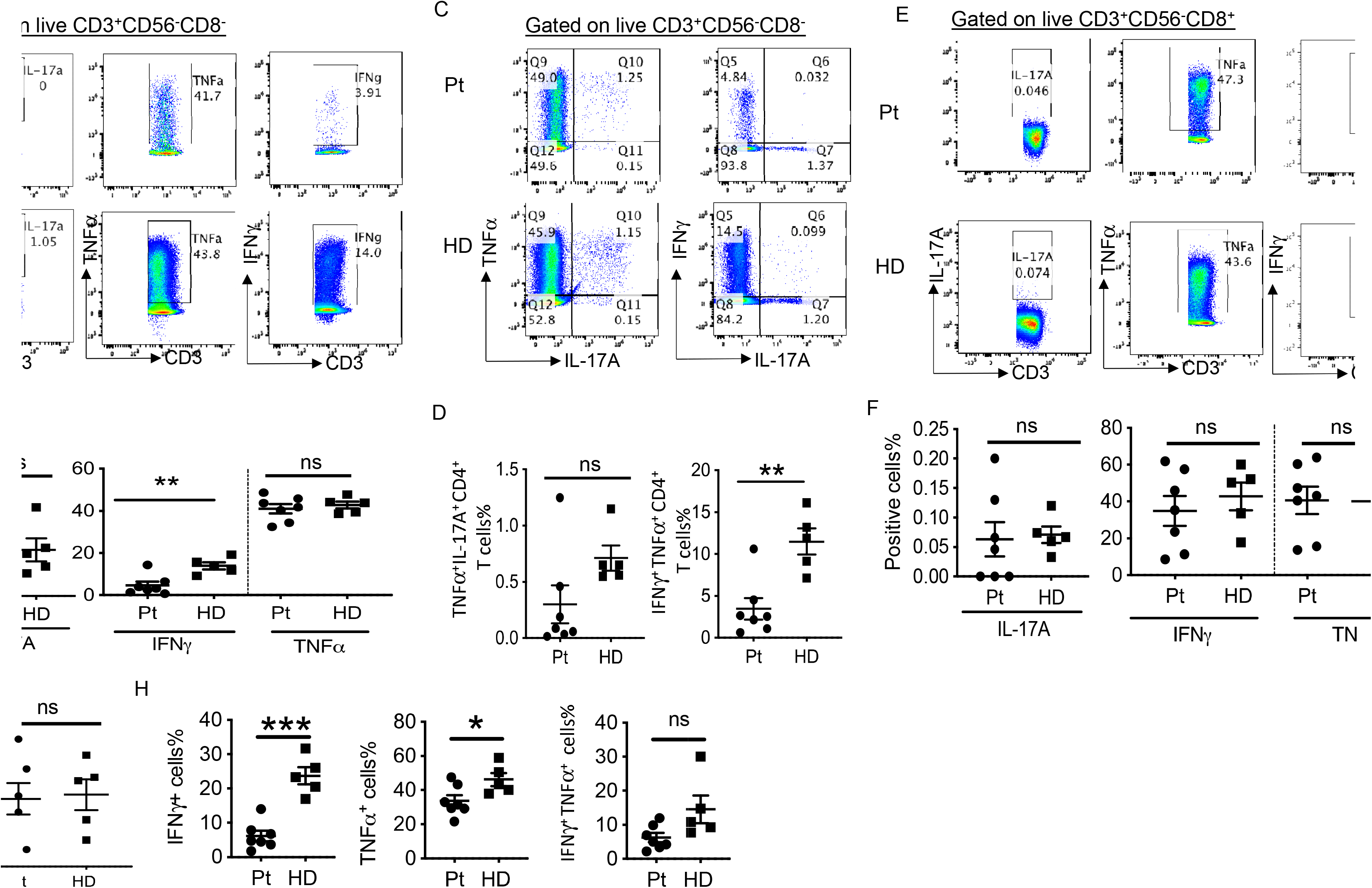
Cellular functionality of T cells in severe COVID-19 cases. (A) Intracellular cytokine staining of CD3^+^CD56^-^CD8^-^ T cells. (B) Summarized data on the frequencies of cytokine-producing CD3^+^CD56^-^CD8^-^ T cells as indicated. (C) Representative FACS plots showing polyfunctional CD3^+^CD56^-^CD8^-^T cells. (D) Summarized data on the frequencies of polyfunctional CD3^+^CD56^-^CD8^-^ T cells as indicated. (E) Intracellular cytokine staining of CD3^+^CD56^-^CD8^+^ T cells. (F) Summarized data on the frequencies of cytokine-producing CD3^+^CD56^-^CD8^+^ T cells as indicated. (G) Summarized data on the frequencies of polyfunctional CD3^+^CD56^-^CD8^+^ T cells as indicated. (H) Summarized data on the frequencies of cytokine-producing CD3^-^cells as indicated. HD, healthy donors (HD#10-14); Pt, patients (n=7). Date are presented as Mean ± SEM. *P<0.05, 0.05<**P<0.001, ***P<0.001. ns, not significant.

Despite the decreased frequencies of CD8^+^ T cells in PBMCs from the patients, these CD8^+^ T cells expressed similar levels of cytokines IFNγ and TNFα as the ones from healthy donors (Figure 4E and 4F). In addition, similar frequencies of IFNγ^+^TNFα^+^ CD8^+^ T cells were found in severe patients and healthy donors (Figure 4G).

We also evaluated capacities of cytokine production in CD3^-^cells, likely NK cells. As shown in Figure 4H, these cells in severe patients expressed significantly reduced levels of IFNγ and TNFα than those in healthy donors. We did not observe significant difference in terms of the frequency of IFNγ^+^ TNFα^+^ cells between these two groups (Figure 4H).

### Cellular immune responses to SARS-CoV-2 in severe COVID-19 subjects

To further measure virus-specific cellular immunity, we treated PBMCs with recombinant NP, S-RBD and S proteins, followed by IFNγ ELISpot analysis. As shown in Figure 5A, the absolute numbers of T cells produced IFNγ in response to anti-CD3 were decreased dramatically in PBMCs from severe patients than those from healthy donors. More strikingly, we did not detect any SARS-CoV-2-specific T cells in all the tested severe patients, whereas one out of ten T cells could secret IFNγ after exposure to NP protein in convalescent individuals (Figure 5B).

**Figure 5.**
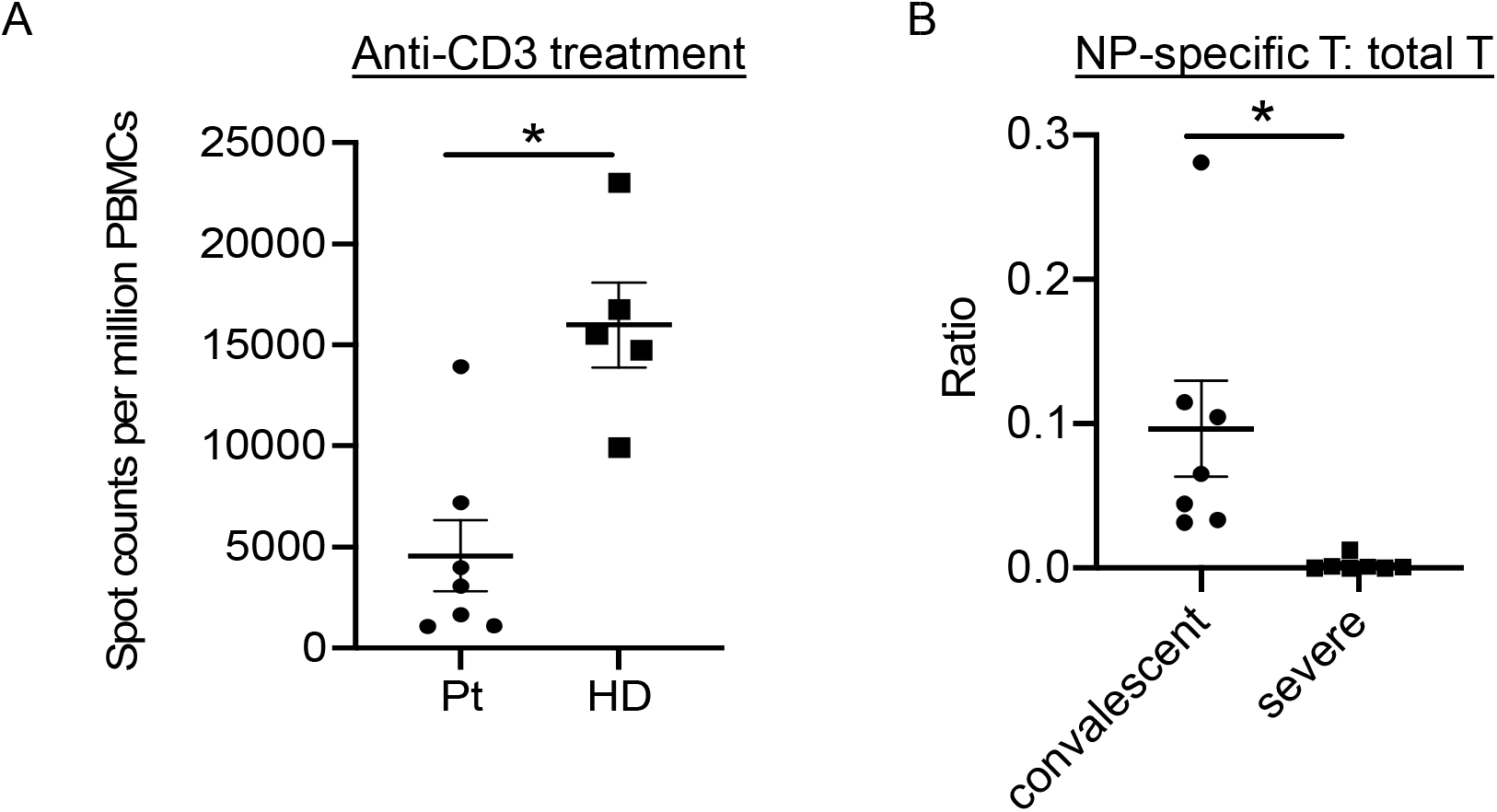
T cell responses to recombinant SARS-CoV-2 proteins in COVID-19 patients. (A) IFNγ ELISpot analysis of COVID-19 patients to anti-CD3 antibody. HD, healthy donors (HD#10-14); Pt, patients (n=7). The experiment with patients was performed in duplicates. (B) The ratios of IFNγ-producing T cells in response to NP or anti-CD3 stimulation in both convalescent (n=14) and severe (n=7) patients. Date are presented as Mean ± SEM. *P<0.05, 0.05<**P<0.001, ***P<0.001.

Thus, T cells in the severe COVID-19 patients failed in developing cellular immunity to SARS-CoV-2.

## Discussion

In this study, we characterized humoral and cellular immunity in severe COVID-19 patients during hospitalization. Although most patients developed anti-SARS-CoV-2 IgG and IgM responses, humoral immune responses varied widely among the patients. The percentages and absolute numbers of both NK cells and CD8^+^ T cells were significantly reduced, accompanied with decreased Th1 cell response in peripheral blood from severe patients. Most notably, we failed in SARS-CoV-2-specific IFNγ production in these patients.

In this study, most severe patients had reduced Th1 cell responses, consistent with the published report (Chen et al., 2020). Another report showed that peripheral blood of a severe COVID-19 patient had a strikingly high number of CCR6^+^ Th17 cells (Xu et al., 2020). In addition, several papers proposed use of therapies directed at Th17 cells and IL-17A signaling in treating COVID-19 patients (De Biasi et al., 2020; Orlov et al., 2020; Wu and Yang, 2020). However, we observed no significant Th17 cell responses in most of the severe patients (5/7) with only one patient showing elevated IL-17A expression. Several possibilities may account for the discrepancy. One is different markers used for defining Th17 cells. The above-mentioned paper used CCR6 on CD4^+^ T cells to define Th17 cells, whereas we used IL-17A expression in CD4^+^ T cells. Another is different patient cohort and disease severity. Sample timing during disease course is another possibility, since T cell responses are dynamic. Given not all the severe COVID-19 patients had increased IL-17A expression, one may not want to treat all patients with IL-17A inhibitors.

5 healthy donors (HD#10-14) with a mean age of 29.4 years were included in our T cell assays, which are not age-matched with severe patients with a mean age of 57 years. Compared to healthy controls, severe patients showed reduced percentages of CD8^+^ T cells and IFNγ-expressing CD4^+^ T cells. Previously, Carr et al found that frequencies of Th1 and CD8^+^ T cells were enhanced with age (Carr et al., 2016). Thus, the differences we observed between healthy controls and severe patients were unlikely caused by age.

In our study, antigens-specific IFNγ expression was not detected in severe patients, in contrast to the report (Weiskopf et al., 2020). We used antigen-mediated IFNγ ELISpot analysis to detect antigen-specific T cells, while the other group used T cell receptor-dependent activation marker assay in measuring the expression of CD137 and CD69. Since AIM assay did not measure effector function by T cells, it is possible that virus-specific T cells are “exhausted”. Different patient cohort and sample timing may account for the different results.

In summary, we detected anti-SARS-CoV-2 antibody responses in most severe cases, while impaired cellular responses were observed in all severe COVID-19 patients. Our results thus provide insight in the pathogenesis of severe COVID-19. They suggest that induction of cellular immunity is vital in controlling SARS-CoV-2 infection, which also has implications in development of an effective vaccine.

## Data Availability

all the data referred to in the manuscript are available.

## ACKNOWLEDGEMENT

We thank Weijin Huang and Jianhui Nie from the National Institutes for Food and Drug Control for sharing the plasmids for pseudovirus package. This work was supported in part by grants from the National Key Research and Development Program of China (2016YFC0906200 to CD, 2016YFC130390 to LN, and 2020YFA0707800 to XW), Natural Science Foundation of China (31991173, 31821003 and 31991170 to CD), Beijing Municipal Science and Technology (Z181100001318007, Z181100006318015 and Z171100000417005 to C.D.), Zhejiang University Foundation (2020XGZX014 to CD), Tsinghua University (to CD) and Science and Technology Development Plan Project of Jilin Province (20200901007SF to F.L.).

## AUTHOR CONTRIBUTIONS

L.N. and C.D. designed the research and analyzed the data. Y.F. and F.C. collected clinical specimens with mild COVID-19; J.L. and H.Z. collected clinical specimens with severe COVID-19; Y.D., X.L. and Q.Y. did most of the experiments at a P3 laboratory. M.C., Y.F., H.Z., Q.Y., Z.G., L.X., P. W., S.J. and D.Y. performed some experiments or prepared key reagents; L.N. and C.D. analyzed the results; L.N. and C.D. wrote the manuscript.

## STAR METHODS

### KEY RESOURCES TABLE

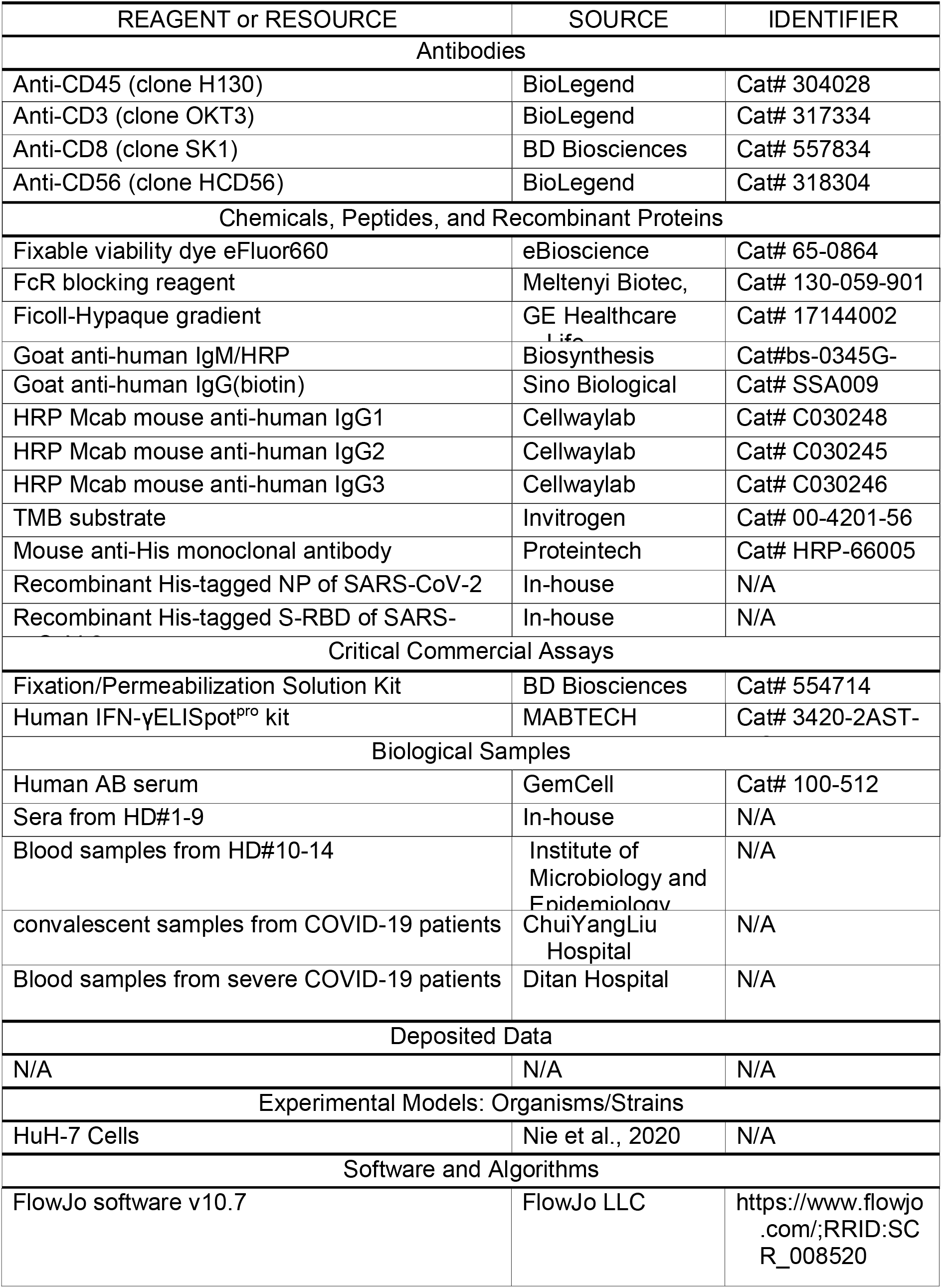

## RESOURCE AVAILABILITY

### Lead Contact

Further information and requests for resources and reagents should be directed to and will be fulfilled by the lead contact, Chen Dong

### Materials Availability

The plasmids (pET28-N-6XHis and pEF1a-S-RBD-6His) generated in this study will be made available on request from the Lead Contact without restriction.

### Data and Code Availability

The study did not generate any unique dataset or code.

## EXPERIMENTAL MODEL AND SUBJECT DETAILS

### COVID-19 patient blood samples

The blood samples of severe COVID-19 patients defined as severe lower respiratory tract infection or pneumonia with fever plus any one of the following: tachypnea, respiratory distress, or oxygen saturation less than 93% on room air according to the guidelines released by the national Health Commission of China were obtained from Ditan hospital in Beijing. All procedures followed were in accordance with the ethical standards of the responsible committee on human experimentation (the institutional review board at Tsinghua University and at Ditan hospital) and with the Helsinki Declaration of 1975, as revised in 2000. All studies were approved by the Medical Ethical Committee at Tsinghua University. Informed consent was obtained from all subjects for being included in the study. All patient data were anonymized before study inclusion. See Table 1 for full patient information, including age, sex, and health status.

### Cell Lines

HuH-7 cells originally taken from a liver tumor in a Japanese male were cultured in DMEM supplemented with 10% FBS. Cells were grown at 37 °C in a 5% CO2 setting.

## METHOD DETAILS

### Expression and Purification of recombinant proteins

The recombinant His-tagged NP of SARS-CoV-2 was expressed in E. coli by a T7 expression system, with 1 mM IPTG induction at 37 ºC for 4 h. The recombinant His-tagged S-RBD (amino acids 319-541) was expressed by a mammalian system in 293F cells. Purified proteins were identified by SDS-PAGE gels and stained with Coomassie blue.

### Isolation of PBMC

PBMCs were isolated from anti-coagulant blood using Ficoll-Hypaque gradients (GE Healthcare Life Sciences, Philadelphia, PA) as previously described (Xie et al., 2018) under the biosafety level 3 facility in AMMS. To isolate PBMCs, blood diluted with PBS, was gently layered over an equal volume of Ficoll in a Falcon tube and centrifuged for 30-40 minutes at 400-500 g without brake. Four layers formed, each containing different cell types. The second layer contained PBMCs. These cells could be gently removed using a Pasteur pipette and added to warm medium or PBS to wash off any remaining platelets. The pelleted cells were then counted and the percentage viability was estimated using Trypan blue staining. Cells were then cryopreserved for future study.

### Anti-SARS-CoV-2 IgG/IgM ELISA

For IgM/IgG testing, 96-well ELISA plates were coated overnight with recombinant NP and S-RBD (100 ng/well). The sera from COVID-19 patients were incubated for 1 h at 37°C. An anti-Human IgG-biotin conjugated monoclonal antibody (Cat. SSA009, Sino Biological Inc., Wayne, PA) and streptavidin-HRP were used at a dilution of 1: 5000 and 1:250, respectively, and anti-human IgM-HRP conjugated monoclonal antibody (Cat. bs-0345G-HRP, Biosynthesis Biotechnology Inc. Beijing, China) was used. The OD value at 450 nm was calculated. The area under the curve (AUC) was calculated by Prism 8 (Graphpad). As a second analytical approach (Grifoni et al., 2020), the serum from one convalescent mild COVID-19 patient was used as a positive control standard. In order to quantify the amount of anti-NP/S-RBD IgG or anti-NP/S-RBD IgM present in each specimen, the positive control standard was run on each plate to calculate antibody titers (relative units) for all samples using non-linear regression interpolations.

### Anti-SARS-CoV-2 IgG1/IgG3 ELISA

For IgG1/IgG2/IgG3 test, 96 well ELISA plates were coated (80 ng/well) overnight with recombinant NP and S-RBD. Plates were washed and the sera from COVID-19 patients were incubated for 1 h at 37°C. After washing, an anti-Human IgG1-HRP conjugated monoclonal antibody (Cat. C030248, BaiaoTong Experiment Center, LY), and an anti-human IgG3-HRP conjugated monoclonal antibody (Cat.C030246, BaiaoTong Experiment Center, LY), all validated by the company for their specificity, were used at a dilution of 1:4000 for 1 h at RT. After washing, TMB substrate solution was added. The OD value at 450 nm was calculated. The area under the curve (AUC) was calculated by Prism 8 (Graphpad).

### Neutralizing antibody assay

Pseudovirus expressing the SARS-CoV-2 S protein was produced. pNL43Luci and GP-pCAGGS were co-transfected into 293T cells. 48 hours later, SARS-CoV-2 pseudovirus-containing supernatants were mixed with at least 6 serially diluted serum samples from the COVID-19 patients at 37°C for 1 hour. Then the mixtures were transferred to 96-well plates containing monolayers of Huh-7 cells (Nie et al., 2020). 3 hours later, the medium was replaced. After incubation for 48 h, the cells were washed, harvested in lysis buffer and analyzed for luciferase activity by the addition of luciferase substrate. Inhibition rate = [1-(the sample group- the cell control group) / (the virus control group-the cell control group)] × 100%. The neutralizing antibody titer (NAT50) were calculated by performing S-fit analysis via Graphpad Prism 7 software.

### Interferon Gamma (IFNγ) ELISpot

IFN-γ-secreting T cells were detected by Human IFNγ ELISpot^pro^ kits (MABTECH AB, Sweden) according to the manufacture protocol. Fresh PBMCs were plated in duplicate at 150k per well and then incubated 48h with 1uM of recombinant proteins. Spots were then counted using an AID ELIspot Reader System (iSpot, AID GmbH). The number of spots was converted into the number of spots per million cells and the mean of duplicate wells plotted.

### FACS staining

PBMCs were washed with PBS plus 2% FBS (Gibco, Grand Island, NY), and then Fc blocking reagent (Meltenyi Biotec, Inc., Auburn, CA) was added followed by a wash with PBS plus 2% FBS. Cells were then incubated for 30 min on ice with anti-CD3 (OKT3) (BioLegend), anti-CD8 (SK1) (BD), anti-CD56 (HCD56) (BioLegend) and live/dead fixable aqua dye (eF660, eBioscience), washed twice with PBS plus 2% FBS and then stored at 4 ° C until acquired by FACS Verse (BD Biosciences, San Jose, CA). Data were analyzed using FlowJo software (Version 10.0.8, Tree Star Inc., Ashland, Or).

## QUANTIFICATION AND STATISTICAL ANALYSIS

Prism 8 software is used for statistical analysis. Student’s t test was performed for two-group analysis. Pearson’s correlation coefficients were calculated. *P* values less than 0.05 were considered to be statistically significant.

## Reference

Carr, E.J., Dooley, J., Garcia-Perez, J.E., Lagou, V., Lee, J.C., Wouters, C., Meyts, I., Goris, A., Boeckxstaens, G., Linterman, M.A., and Liston, A. (2016). The cellular composition of the human immune system is shaped by age and cohabitation. Nat Immunol 17, 461–468.

Chen, G., Wu, D., Guo, W., Cao, Y., Huang, D., Wang, H., Wang, T., Zhang, X., Chen, H., Yu, H., et al. (2020). Clinical and immunological features of severe and moderate coronavirus disease 2019. J Clin Invest 130, 2620–2629.

De Biasi, S., Meschiari, M., Gibellini, L., Bellinazzi, C., Borella, R., Fidanza, L., Gozzi, L., Iannone, A., Lo Tartaro, D., Mattioli, M., et al. (2020). Marked T cell activation, senescence, exhaustion and skewing towards TH17 in patients with COVID-19 pneumonia. Nat Commun 11, 3434.

Di Pierro, F., Bertuccioli, A., and Cavecchia, I. (2020). Possible therapeutic role of a highly standardized mixture of active compounds derived from cultured Lentinula edodes mycelia (AHCC) in patients infected with 2019 novel coronavirus. Minerva Gastroenterol Dietol.

Grifoni, A., Weiskopf, D., Ramirez, S.I., Mateus, J., Dan, J.M., Moderbacher, C.R., Rawlings, S.A., Sutherland, A., Premkumar, L., Jadi, R.S., et al. (2020). Targets of T Cell Responses to SARS-CoV-2 Coronavirus in Humans with COVID-19 Disease and Unexposed Individuals. Cell.

Gruszecka, J., and Filip, R. (2020). Preliminary information on prevention of infections caused by SARS-COV-2 virus in endoscopic laboratories. Ann Agric Environ Med 27, 171–174.

Lynch, K.L., Whitman, J.D., Lacanienta, N.P., Beckerdite, E.W., Kastner, S.A., Shy, B.R., Goldgof, G.M., Levine, A.G., Bapat, S.P., Stramer, S.L., et al. (2020). Magnitude and kinetics of anti-SARS-CoV-2 antibody responses and their relationship to disease severity. Clin Infect Dis.

Ni, L., Ye, F., Cheng, M.L., Feng, Y., Deng, Y.Q., Zhao, H., Wei, P., Ge, J., Gou, M., Li, X., et al. (2020). Detection of SARS-CoV-2-Specific Humoral and Cellular Immunity in COVID-19 Convalescent Individuals. Immunity 52, 971–977 e973.

Nie, J., Li, Q., Wu, J., Zhao, C., Hao, H., Liu, H., Zhang, L., Nie, L., Qin, H., Wang, M., et al. (2020). Establishment and validation of a pseudovirus neutralization assay for SARS-CoV-2. Emerg Microbes Infect 9, 680–686.

Orlov, M., Wander, P.L., Morrell, E.D., Mikacenic, C., and Wurfel, M.M. (2020). A Case for Targeting Th17 Cells and IL-17A in SARS-CoV-2 Infections. J Immunol.

Wang, C., Horby, P.W., Hayden, F.G., and Gao, G.F. (2020a). A novel coronavirus outbreak of global health concern. Lancet 395, 470–473.

Wang, Y., Zhang, L., Sang, L., Ye, F., Ruan, S., Zhong, B., Song, T., Alshukairi, A.N., Chen, R., Zhang, Z., et al. (2020b). Kinetics of viral load and antibody response in relation to COVID-19 severity. J Clin Invest.

Weiskopf, D., Schmitz, K.S., Raadsen, M.P., Grifoni, A., Okba, N.M.A., Endeman, H., van den Akker, J.P.C., Molenkamp, R., Koopmans, M.P.G., van Gorp, E.C.M., et al. (2020). Phenotype and kinetics of SARS-CoV-2-specific T cells in COVID-19 patients with acute respiratory distress syndrome. Sci Immunol 5.

Wu, D., and Yang, X.O. (2020). TH17 responses in cytokine storm of COVID-19: An emerging target of JAK2 inhibitor Fedratinib. J Microbiol Immunol Infect 53, 368–370.

Xie, S., Huang, J., Qiao, Q., Zang, W., Hong, S., Tan, H., Dong, C., Yang, Z., and Ni, L. (2018). Expression of the inhibitory B7 family molecule VISTA in human colorectal carcinoma tumors. Cancer Immunol Immunother 67, 1685–1694.

Xu, Z., Shi, L., Wang, Y., Zhang, J., Huang, L., Zhang, C., Liu, S., Zhao, P., Liu, H., Zhu, L., et al. (2020). Pathological findings of COVID-19 associated with acute respiratory distress syndrome. Lancet Respir Med 8, 420–422.

Zheng, H.Y., Zhang, M., Yang, C.X., Zhang, N., Wang, X.C., Yang, X.P., Dong, X.Q., and Zheng, Y.T. (2020). Elevated exhaustion levels and reduced functional diversity of T cells in peripheral blood may predict severe progression in COVID-19 patients. Cell Mol Immunol 17, 541–543.

